# Trends and factors associated with diabetes screening, diagnosis and treatment in Peru, 2014-2024

**DOI:** 10.64898/2025.12.18.25342639

**Authors:** Wilmer Cristobal Guzman-Vilca, Maricela Curisinche-Rojas, Gilmer Solis-Sánchez, Raúl Timaná-Ruiz

## Abstract

**Background:** Timely access to screening, diagnosis, and treatment services is essential to prevent complications among people with diabetes. We evaluated trends and factors associated with self-reported diabetes screening, diagnosis, and treatment among Peruvian adults aged 18 years and older from 2014 to 2024.

**Methods:** We analyzed data from the Health Questionnaire of the Demographic and Family Health Survey (ENDES). We used information on diabetes screening, diagnosis, and treatment, along with socio-demographic and clinical covariates. We fitted generalized linear models of the Poisson family with log link and robust variance, and performed Joinpoint regression to assess temporal trends.

**Results:** The prevalence of screening increased from 24% to 31% (APC: 1%; p=0.483). Higher screening was associated with being female, having higher educational attainment, not being single, belonging to higher wealth quintiles, being insured, and having hypertension. In contrast, smoking and living in rural areas or the Highlands were associated with lower screening. The prevalence of diagnosis increased from 3% to 5% (APC: 7%; p<0.001). Higher diagnosis was associated with being female, not being single, higher wealth, being insured, and living in the Rainforest, while lower diagnosis was associated with higher education and living in rural areas or the Highlands. Among individuals with diabetes, treatment prevalence increased from 65% to 74% between 2014 and 2019 (APC: 2%; p=0.014), but decreased to 63% in 2024 (APC: −2%; p<0.001). Higher treatment was associated with being married/widowed/divorced, being insured, higher wealth, and having hypertension. Lower treatment was associated with higher education and living in the Highlands.

**Conclusions:** Although the prevalences of diabetes screening and diagnosis have increased over the past 11 years, treatment has declined in the last five. Socio-demographic and clinical inequalities persist across all three indicators and should be considered in the planning of diabetes health services in Peru.

## INTRODUCTION

Diabetes is a chronic condition characterized by hyperglicemia [1]. According to the World Health Organization (WHO), the crude prevalence of diabetes among adults aged 18 years and older was 14.8% in 2022, and 14.1% when standardized by age and sex [2]. Moreover, the number of individuals living with diabetes is projected to increase from 537 million in 2021 to 643 million in 2045 [3], with the majority of cases occurring in low- and middle-income countries (LMICs) [4]. Populations at higher risk include adults with overweight or obesity, those with a family history of diabetes, and individuals from high-risk ethnic groups such as Latin Americans [5].

Regular access to screening services enables timely identification of individuals with diabetes. Once diagnosed, patients can receive appropriate treatment and follow-up from healthcare professionals to achieve adequate glycemic control. Failure to ensure this continuum of care increases the risk of both macrovascular and microvascular complications [1]. Factors influencing the development of complications include adherence to healthy lifestyles, diabetes duration, and pharmacological management [6]. These determinants depend largely on the availability and quality of healthcare services for individuals with diabetes. Nevertheless, diagnosis and treatment rates in LMICs remain low [7].

A cross-sectional study conducted in LMICs reported that among all individuals identified with diabetes, 44% had previously received a diagnosis and only 38% of those diagnosed were receiving treatment [8]. In Peru, one study found that in 2020, 28% of individuals aged 15 years and older had been screened for diabetes within the previous 12 months based on data from the Demographic and Family Health Survey (ENDES in Spanish) [9]. Additionally, the National Institute of Statistics and Informatics (INEI in Spanish), using ENDES 2024 data, reported that 5% of the population aged 15 years or older self-reported a diagnosis of diabetes, and 73% of those with diabetes had received treatment in the previous 12 months [10]. Although these studies are informative, their results were only for one year, which limits the evaluation of trends in screening, diagnosis and treatment for people with diabetes and thus, whether it has been improvement on these indicators is unknown.

Multiple factors influence access to screening, diagnosis, and treatment services for diabetes. A systematic review found that key reasons for not attending the National Health Service screening services in the United Kingdom included the lack of awareness about screening availability and purpose, time constraints (e.g., work obligations), and difficulties in accessing services [11]. Similarly, a study from Singapore reported that sociodemographic factors associated with regular diabetes screening included ethnicity, older age, higher education, and monthly personal income [12]. In China, factors associated with higher odds of diabetes diagnosis included older age, family history of diabetes, obesity, and elevated levels of systolic blood pressure, triglycerides, and high-density lipoprotein cholesterol [13]. Regarding treatment, a cross-sectional study spanning 55 LMICs found that being female, older age, and higher body mass index (BMI), education, and income were associated with greater likelihood of receiving diabetes treatment [7].

To date, there has been no comprehensive analysis of access to diabetes screening, diagnosis, and treatment services in Peru. Although INEI provides annual descriptive estimates of self-reported diagnosis and treatment [10], these reports do not include diabetes screening, do not analyze temporal trends rigorously, and do not examine variation across sociodemographic, clinical, or regional characteristics.

Given this context and the limited evidence on the use of health services for diabetes in Peru, this study aimed to assess trends and factors associated with self-reported diabetes screening, diagnosis, and treatment among adults aged 18 years and older in Peru between 2014 and 2024.

## MATERIALS AND METHODS

### Study design

We conducted a an observational, analytical, and longitudinal design, using individual-level data from the ENDES between 2014 to 2024. ENDES is conducted annually by the National Institute of Statistics and Informatics (INEI) and targets households, children, adolescents, adults, and older adults in Peru. The study population includes private households and their usual residents or non-usual residents who slept in the dwelling the night before the interview. Among its components, ENDES administers a Health Questionnaire to individuals aged 15 years and older, selecting one respondent per household. The ENDES sampling design is probabilistic, two-stage, stratified, and independent, allowing for inference at the departmental level and by urban and rural areas [14].

### Study population

The study population included all records of individuals aged ≥18 years who responded to the Health Questionnaire (inclusion criteria). No sampling design was applied, as the analysis included the full set of available observations, excluding those with missing data in any variable of interest (exclusion criteria). This population allowed the estimation of the prevalence of diagnosed diabetes (PDD). Two additional subpopulations were constructed to estimate: i) prevalence of diabetes screening (PDS), excluding individuals with previously diagnosed diabetes; and ii) prevalence of diabetes treatment (PDT), including only respondents with a prior diagnosis of diabetes.

### Variables

PDS, PDD, and PDT, were defined dichotomously (Yes/No) based on self-reported data on having undergone diabetes screening, received a diabetes diagnosis, or received diabetes treatment in the last 12 months. “Don’t know” responses were coded as “No”.

Independent sociodemographic and clinical variables were selected based on prior literature: age [12,15,16], sex [10,17,18], ethnicity [12,15], education level [12,19], marital status [12,16,19], employment status (based on economic activity during the previous week) [12,16,20], wealth index (quintiles) [21,22], health insurance coverage and type (*Seguro Integral de Salud* (SIS), EsSalud, Armed Forces and Police, Health Provider Entity and Private) [16,19,23], area of residence, natural region, region, hypertension [17], daily smoking [24], and body mass index (BMI) [8,17,25] calculated from measured weight and height.

Data were collected by INEI as part of its routine survey operations, during which ENDES questionnaires are administered through in-person interviews conducted by trained fieldworkers [26]. ENDES instruments were developed in accordance with standardized indicators from the DHS Program [27]. Data were downloaded from INEI’s public microdata platform (https://proyectos.inei.gob.pe/microdatos/).

### Statistical analysis

For each study year (2014–2024), we downloaded on September 19^th^, 2025, the following databases: RECH0, RECH1, RECH4, RECH5, RECH23, and CSALUD01. These files were merged within each year using the household and individual identifiers via the *merge* command in STATA, and annual datasets were subsequently combined into a single multiyear database using the *append* command in STATA.

All analyses accounted for the complex survey design using the *svyset* command with the “singleunit(centered)” option, a corrected sampling weight, and a multiyear adjustment by creating a combined strata-year variable [28].

Descriptive analyses of categorical variables included unweighted absolute and relative frequencies (%), as well as weighted percentages (w%) with 95% confidence intervals (95% CI) and coefficients of variation. For numerical variables, we estimated unweighted means and standard deviations, along with weighted means and their 95% CIs. For bivariate analysis, PDS, PDD and PDT were compared using the second-order Rao–Scott chi-square test, while differences in weighted means were assessed using adjusted Wald tests.

Multivariable analyses were performed using generalized linear models (GLM) with Poisson family, log link function, and robust variance, estimating prevalence ratios (PR). Crude models were fitted for each dependent variable, and variables with *p*<0.20 in crude models were included in adjusted models. Variables with a variance inflation factor (VIF) >10 were excluded to avoid multicollinearity [29].

Trend analyses were conducted using Joinpoint regression to identify significant changes in the weighted percentages of PDS, PDD, and PDT, estimating annual percent changes (APC) for each indicator. Trends were also evaluated stratified by variables significant in adjusted models.

Data cleaning, processing and descriptive, bivariate, and multivariable analyses were performed using STATA v19.5 (Stata Corporation, College Station, Texas, USA), and trend analyses were conducted using Joinpoint Desktop v5.1.0.0 (Statistical Research and Applications Branch, National Cancer Institute). A p-value of < 0.05 was considered statistically significant for all analyses.

### Ethics

Our study involves a secondary-data analysis and no human subjects were directly involved in the project, entailing thus a minimal risk. The study was approved by the Peru’s National Institute of Health Research Ethics Committee with the code OI-019-25.

## RESULTS

Following the construction of the database, of the 372,750 participants initially included in the Health Questionnaires of the 2014–2024 ENDES, 321,769 met the eligibility criteria and were included in the analysis. The distribution of these participants by survey year is shown in Figure 1.

**Figure 1.**
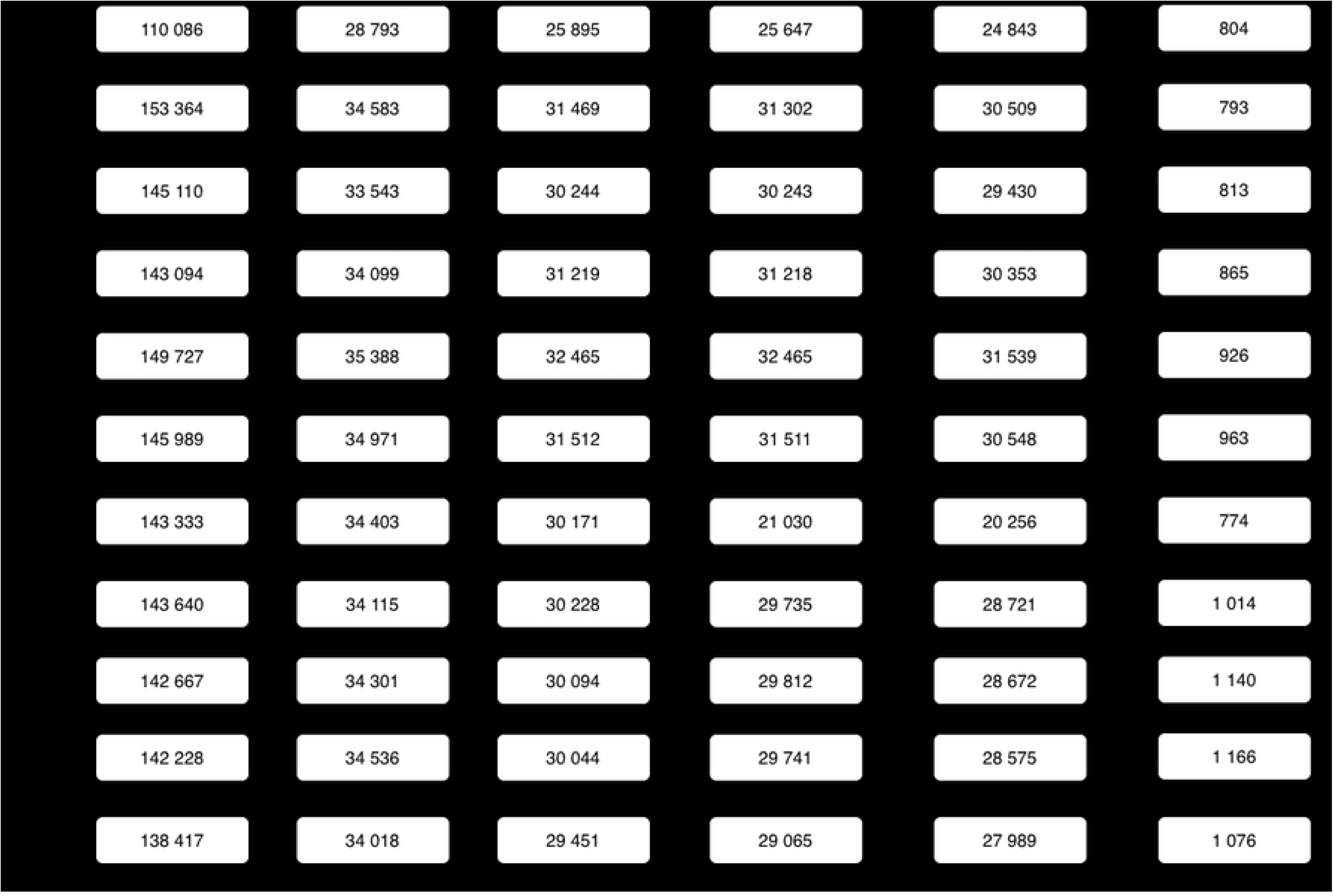
Flowchart of participants included in the analysis by year.

The characteristics of the study population for 2014 and 2024 are presented in Table 1, while detailed information for each year is available in the Supplementary Material (Table S1). The mean age of the population was 42.6 years (95% CI: 42.5–42.7) and remained relatively stable throughout the study period. The most frequent age group was individuals under 40 years, decreasing from 49.0% in 2014 to 46.5% in 2024. The proportion of women remained consistently higher than that of men (52.4% vs. 47.6% in 2024).

**Table 1.**
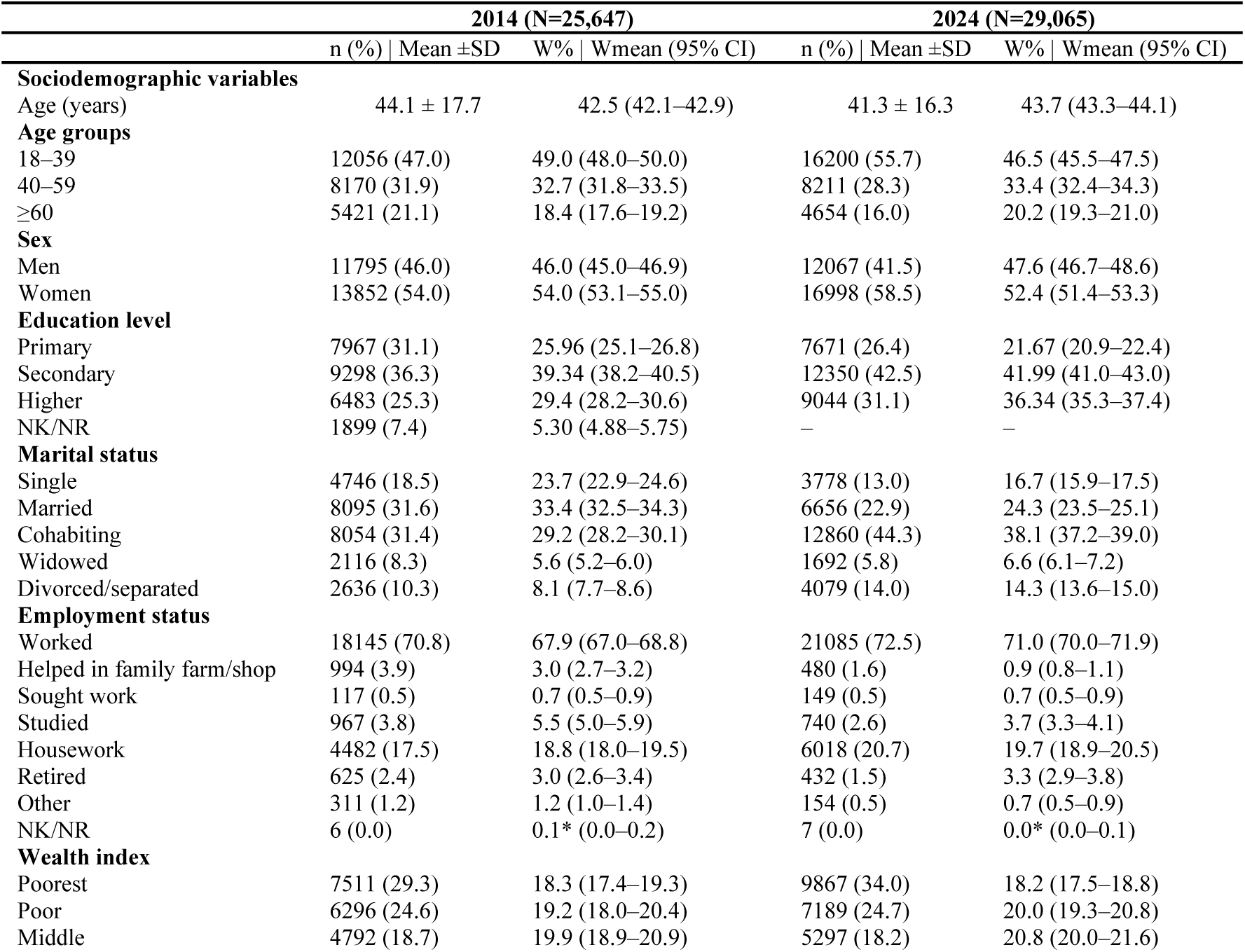
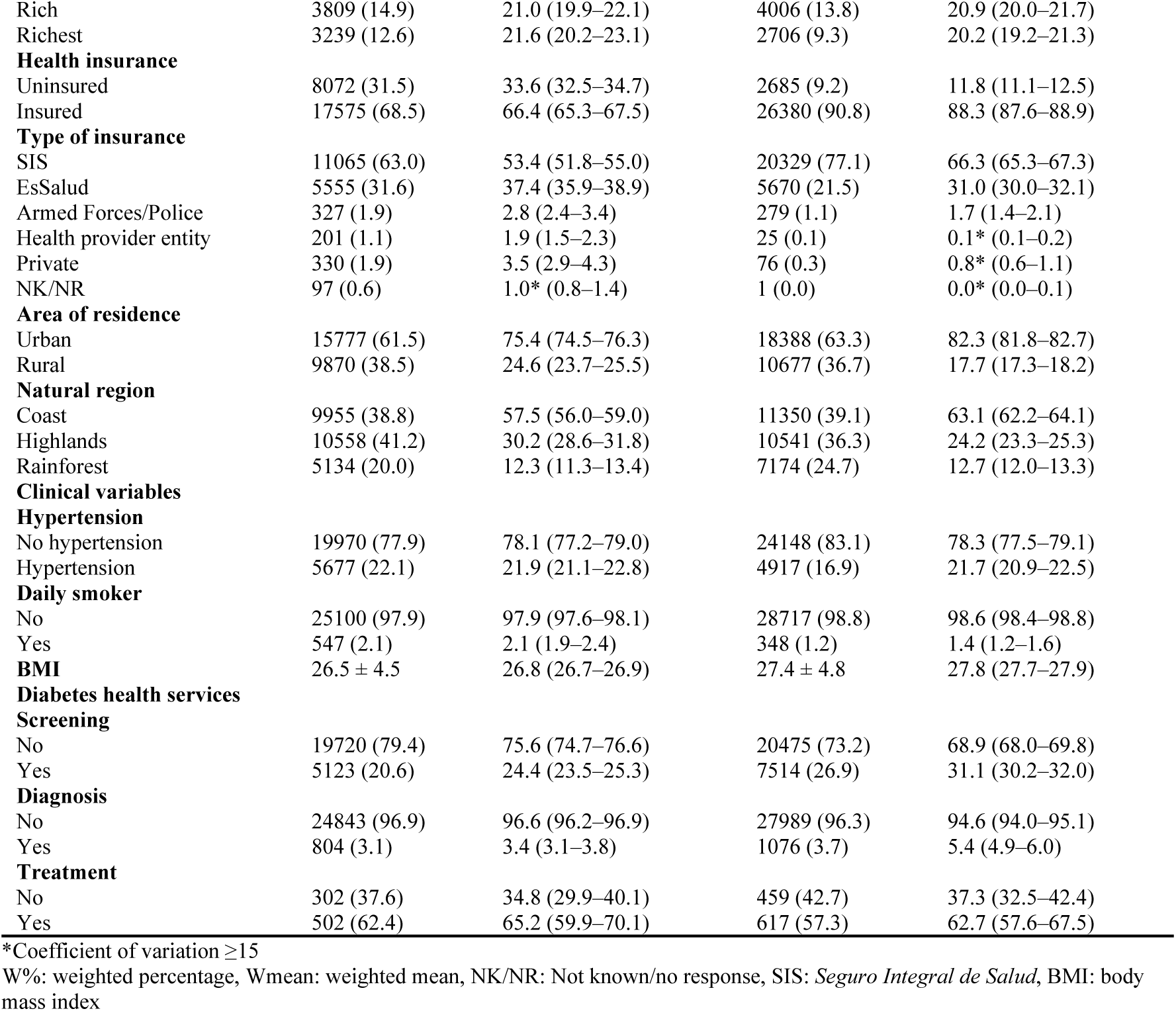
Unweighted and weighted characteristics of the study population in 2014 and 2024.

|  | 2014 (N=25,647) |  |  | 2024 (N=29,065) |  |
| --- | --- | --- | --- | --- | --- |
|  | n (%) \| Mean ±SD | W% \| Wmean (95% CI) |  | n (%) \| Mean ±SD | W% \| Wmean (95% CI) |
| **Sociodemographic variables** |  |  |  |  |  |
| Age (years) | 44.1 ± 17.7 | 42.5 (42.1–42.9) |  | 41.3 ± 16.3 | 43.7 (43.3–44.1) |
| **Age groups** |  |  |  |  |  |
| 18–39 | 12056 (47.0) | 49.0 (48.0–50.0) |  | 16200 (55.7) | 46.5 (45.5–47.5) |
| 40–59 | 8170 (31.9) | 32.7 (31.8–33.5) |  | 8211 (28.3) | 33.4 (32.4–34.3) |
| ≥60 | 5421 (21.1) | 18.4 (17.6–19.2) |  | 4654 (16.0) | 20.2 (19.3–21.0) |
| **Sex** |  |  |  |  |  |
| Men | 11795 (46.0) | 46.0 (45.0–46.9) |  | 12067 (41.5) | 47.6 (46.7–48.6) |
| Women | 13852 (54.0) | 54.0 (53.1–55.0) |  | 16998 (58.5) | 52.4 (51.4–53.3) |
| **Education level** |  |  |  |  |  |
| Primary | 7967 (31.1) | 25.96 (25.1–26.8) |  | 7671 (26.4) | 21.67 (20.9–22.4) |
| Secondary | 9298 (36.3) | 39.34 (38.2–40.5) |  | 12350 (42.5) | 41.99 (41.0–43.0) |
| Higher | 6483 (25.3) | 29.4 (28.2–30.6) |  | 9044 (31.1) | 36.34 (35.3–37.4) |
| NK/NR | 1899 (7.4) | 5.30 (4.88–5.75) |  | – | – |
| **Marital status** |  |  |  |  |  |
| Single | 4746 (18.5) | 23.7 (22.9–24.6) |  | 3778 (13.0) | 16.7 (15.9–17.5) |
| Married | 8095 (31.6) | 33.4 (32.5–34.3) |  | 6656 (22.9) | 24.3 (23.5–25.1) |
| Cohabiting | 8054 (31.4) | 29.2 (28.2–30.1) |  | 12860 (44.3) | 38.1 (37.2–39.0) |
| Widowed | 2116 (8.3) | 5.6 (5.2–6.0) |  | 1692 (5.8) | 6.6 (6.1–7.2) |
| Divorced/separated | 2636 (10.3) | 8.1 (7.7–8.6) |  | 4079 (14.0) | 14.3 (13.6–15.0) |
| **Employment status** |  |  |  |  |  |
| Worked | 18145 (70.8) | 67.9 (67.0–68.8) |  | 21085 (72.5) | 71.0 (70.0–71.9) |
| Helped in family farm/shop | 994 (3.9) | 3.0 (2.7–3.2) |  | 480 (1.6) | 0.9 (0.8–1.1) |
| Sought work | 117 (0.5) | 0.7 (0.5–0.9) |  | 149 (0.5) | 0.7 (0.5–0.9) |
| Studied | 967 (3.8) | 5.5 (5.0–5.9) |  | 740 (2.6) | 3.7 (3.3–4.1) |
| Housework | 4482 (17.5) | 18.8 (18.0–19.5) |  | 6018 (20.7) | 19.7 (18.9–20.5) |
| Retired | 625 (2.4) | 3.0 (2.6–3.4) |  | 432 (1.5) | 3.3 (2.9–3.8) |
| Other | 311 (1.2) | 1.2 (1.0–1.4) |  | 154 (0.5) | 0.7 (0.5–0.9) |
| NK/NR | 6 (0.0) | 0.1* (0.0–0.2) |  | 7 (0.0) | 0.0* (0.0–0.1) |
| **Wealth index** |  |  |  |  |  |
| Poorest | 7511 (29.3) | 18.3 (17.4–19.3) |  | 9867 (34.0) | 18.2 (17.5–18.8) |
| Poor | 6296 (24.6) | 19.2 (18.0–20.4) |  | 7189 (24.7) | 20.0 (19.3–20.8) |
| Middle | 4792 (18.7) | 19.9 (18.9–20.9) |  | 5297 (18.2) | 20.8 (20.0–21.6) |

|  |  |  |  |  |
| --- | --- | --- | --- | --- |
| Rich | 3809 (14.9) | 21.0 (19.9–22.1) | 4006 (13.8) | 20.9 (20.0–21.7) |
| Richest | 3239 (12.6) | 21.6 (20.2–23.1) | 2706 (9.3) | 20.2 (19.2–21.3) |
| **Health insurance** |  |  |  |  |
| Uninsured | 8072 (31.5) | 33.6 (32.5–34.7) | 2685 (9.2) | 11.8 (11.1–12.5) |
| Insured | 17575 (68.5) | 66.4 (65.3–67.5) | 26380 (90.8) | 88.3 (87.6–88.9) |
| **Type of insurance** |  |  |  |  |
| SIS | 11065 (63.0) | 53.4 (51.8–55.0) | 20329 (77.1) | 66.3 (65.3–67.3) |
| EsSalud | 5555 (31.6) | 37.4 (35.9–38.9) | 5670 (21.5) | 31.0 (30.0–32.1) |
| Armed Forces/Police | 327 (1.9) | 2.8 (2.4–3.4) | 279 (1.1) | 1.7 (1.4–2.1) |
| Health provider entity | 201 (1.1) | 1.9 (1.5–2.3) | 25 (0.1) | 0.1* (0.1–0.2) |
| Private | 330 (1.9) | 3.5 (2.9–4.3) | 76 (0.3) | 0.8* (0.6–1.1) |
| NK/NR | 97 (0.6) | 1.0* (0.8–1.4) | 1 (0.0) | 0.0* (0.0–0.1) |
| **Area of residence** |  |  |  |  |
| Urban | 15777 (61.5) | 75.4 (74.5–76.3) | 18388 (63.3) | 82.3 (81.8–82.7) |
| Rural | 9870 (38.5) | 24.6 (23.7–25.5) | 10677 (36.7) | 17.7 (17.3–18.2) |
| **Natural region** |  |  |  |  |
| Coast | 9955 (38.8) | 57.5 (56.0–59.0) | 11350 (39.1) | 63.1 (62.2–64.1) |
| Highlands | 10558 (41.2) | 30.2 (28.6–31.8) | 10541 (36.3) | 24.2 (23.3–25.3) |
| Rainforest | 5134 (20.0) | 12.3 (11.3–13.4) | 7174 (24.7) | 12.7 (12.0–13.3) |
| **Clinical variables** |  |  |  |  |
| **Hypertension** |  |  |  |  |
| No hypertension | 19970 (77.9) | 78.1 (77.2–79.0) | 24148 (83.1) | 78.3 (77.5–79.1) |
| Hypertension | 5677 (22.1) | 21.9 (21.1–22.8) | 4917 (16.9) | 21.7 (20.9–22.5) |
| **Daily smoker** |  |  |  |  |
| No | 25100 (97.9) | 97.9 (97.6–98.1) | 28717 (98.8) | 98.6 (98.4–98.8) |
| Yes | 547 (2.1) | 2.1 (1.9–2.4) | 348 (1.2) | 1.4 (1.2–1.6) |
| **BMI** |  |  |  |  |
|  | 26.5 ± 4.5 | 26.8 (26.7–26.9) | 27.4 ± 4.8 | 27.8 (27.7–27.9) |
| **Diabetes health services** |  |  |  |  |
| **Screening** |  |  |  |  |
| No | 19720 (79.4) | 75.6 (74.7–76.6) | 20475 (73.2) | 68.9 (68.0–69.8) |
| Yes | 5123 (20.6) | 24.4 (23.5–25.3) | 7514 (26.9) | 31.1 (30.2–32.0) |
| **Diagnosis** |  |  |  |  |
| No | 24843 (96.9) | 96.6 (96.2–96.9) | 27989 (96.3) | 94.6 (94.0–95.1) |
| Yes | 804 (3.1) | 3.4 (3.1–3.8) | 1076 (3.7) | 5.4 (4.9–6.0) |
| **Treatment** |  |  |  |  |
| No | 302 (37.6) | 34.8 (29.9–40.1) | 459 (42.7) | 37.3 (32.5–42.4) |
| Yes | 502 (62.4) | 65.2 (59.9–70.1) | 617 (57.3) | 62.7 (57.6–67.5) |

\*Coefficient of variation $\geq 15$
W%: weighted percentage, Wmean: weighted mean, NK/NR: Not known/no response, SIS: *Seguro Integral de Salud*, BMI: body mass index

Regarding educational attainment, the proportion of individuals with completed higher education increased from 29.4% in 2014 to 36.3% in 2024. No major variations were observed across the study period in terms of employment status or wealth quintiles. By marital status, the proportion of married individuals decreased from 33.4% in 2014 to 24.3% in 2024, while the proportion of cohabiting individuals increased from 29.2% to 38.1%. During this period, health insurance coverage rose from 66.4% to 88.3%.

In 2024, 82.3% of the population resided in urban areas and 63.1% in the Coastal region, both proportions higher than those observed in 2014. Although the prevalence of hypertension exhibited minor fluctuations over time, it remained virtually unchanged between 2014 and 2024 (21.9% vs. 21.7%). The proportion of daily smokers decreased from 2.1% in 2014 to 1.4% in 2024, while mean BMI increased from 26.8 kg/m² to 27.8 kg/m² (Table 1).

PDS increased steadily from 22.5% in 2015 to 29.7% in 2019, declined during 2020–2021 (19.8%), and increased again toward 2024 (31.1%). PDD rose from 3.0% in 2015 to 5.7% in 2023, followed by a slight decrease in 2024 (5.4%). Among individuals with diabetes, PDT increased from 65.2% in 2014 to 74.4% in 2019, decreased in 2020 (66.2%), remained stable until 2023 (66.5%), and declined in 2024 (62.7%) (Table 1).

PDS, PDD, and PDT according to population characteristics are presented in Table 2 for 2014 and 2024, with year-specific estimates available in Supplementary Material (Table S2). Across the study period, mean age was significantly higher among individuals who had undergone screening, received a diagnosis, or received treatment for diabetes compared to those who had not (p<0.05 for 2014–2024). By sex, although PDS and PDD were consistently higher among women, the difference in diagnosis was not significant in 2014, 2016–2018, or 2020–2022.

**Table 2.**
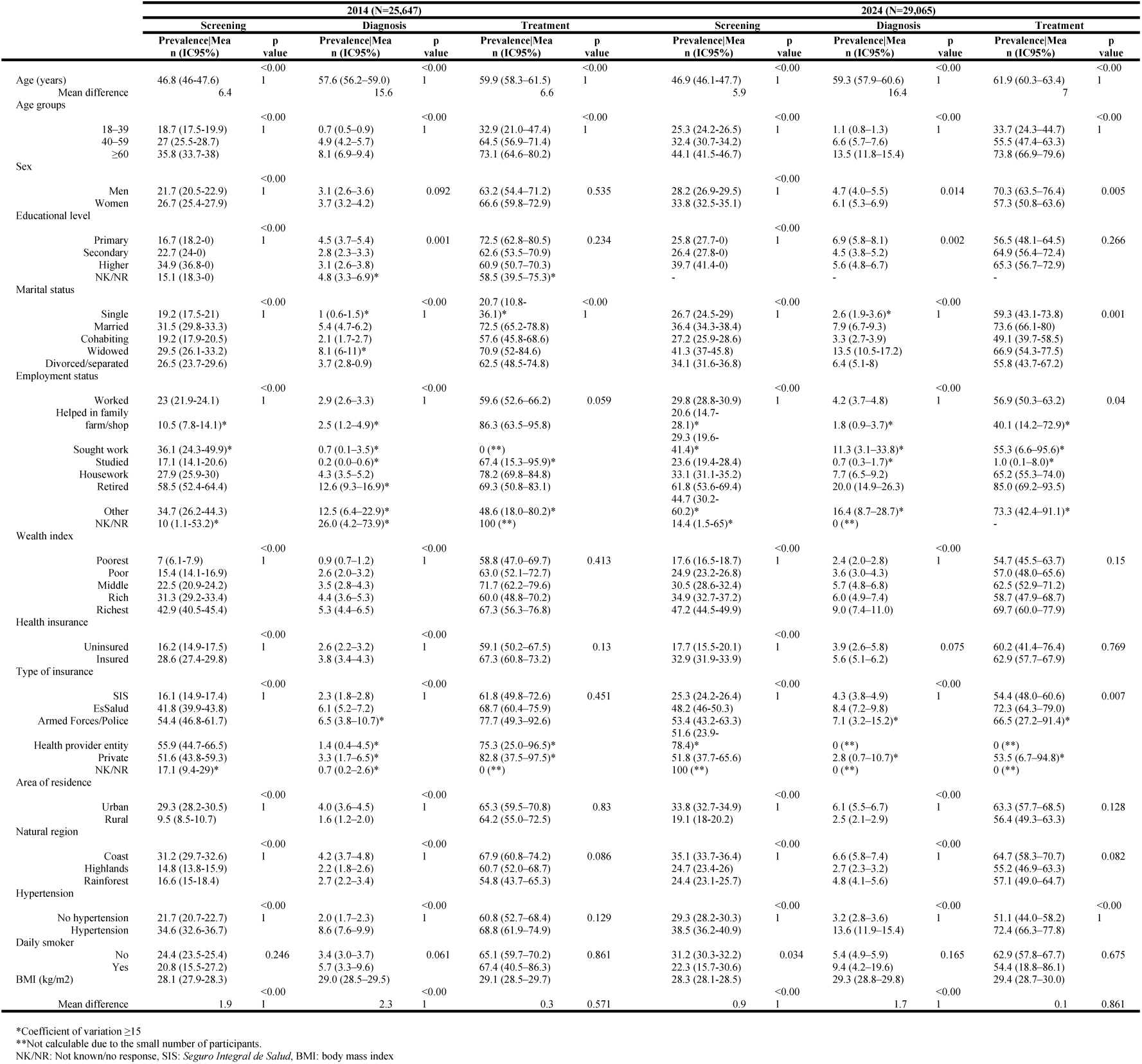
Weighted distribution of screening, diagnosis and treatment of diabetes according to the characteristics of the study population in 2014 and 2024.

Although PDT was slightly higher among women during 2014–2019 (p<0.05 in 2015 and 2018), it became higher among men from 2020 onward (p<0.05 in 2024). While PDS increased with higher educational attainment, PDD was generally higher among those with primary education or less. Since 2017, PDT showed no significant differences by educational level. Across the study period, the highest prevalence of screening was observed among widowed and married individuals, diagnosis was highest among widowed individuals, and treatment was highest among widowed and married individuals (Table 2 and S2).

By employment status, PDS, PDD, and PDT were higher among retirees throughout the study period, except for PDT in 2021, where no significant differences were observed. PDS and PDD were higher among individuals from wealthier households, decreasing with lower wealth quintiles across the entire period. A similar pattern was observed for

PDT from 2015 onward, although differences were generally not significant (p<0.05 in 2016, 2018, and 2020). PDS and PDD were consistently higher among individuals with health insurance. PDT was also higher among insured individuals throughout the study period, except in 2016 (Table 2 and S2).

Across the study period, PDS (p<0.05 for 2014–2024), PDD (p<0.05 for 2014–2024), and PDT (p<0.05 in 2015 and 2019–2023) were higher among individuals from urban areas and the Coastal region (Table 2).

Individuals with hypertension had higher PDS and PDD throughout the study period, and higher PDT from 2018 onward. No significant differences were observed between smokers and non-smokers for PDS and PDT, except for screening in 2023 and diagnosis in 2017, where prevalence was higher among smokers. Although no consistent differences were observed for PDT, it was higher among non-smokers in 2017 and 2020 and higher among smokers in 2021. Across the study period, mean BMI was higher among individuals who had been screened or diagnosed, but it did not differ according to treatment status. Differences in PDS, PDD, and PDT by ethnicity and region are presented in Supplementary Tables S3 and S4.

### Factors associated with diabetes screening, diagnosis, and treatment

In the adjusted model for factors associated with PDS, screening was significantly higher among women, individuals with higher educational attainment, and those with higher wealth index (p<0.05). Screening was also higher among individuals who were not single, with the strongest association observed for widowed individuals. Having health insurance was associated with a 75% higher PDS in the crude model, decreasing to 69% in the adjusted model (p<0.05). Living in rural areas (vs. urban) and in the Highlands (vs. Coast) was associated with 8% and 14% lower PDS, respectively (p<0.05). Hypertension and smoking were associated with higher PDS, as was higher BMI in the crude model (p<0.05). Age and BMI were not included in the adjusted model due to multicollinearity (Table 3).

**Table 3.** Factors associated with PDS, PDD and PDT in 2014-2024.

|  |  | PDS |  |  |  | PDD |  |  |  | PDT |  |  |  |
| --- | --- | --- | --- | --- | --- | --- | --- | --- | --- | --- | --- | --- | --- |
|  |  | Crude PR (95% CI) | p value | Adjusted PR (95% CI) | p value | Crude PR (95% CI) | p value | Adjusted PR (95% CI) | p value | Crude PR (95% CI) | p value | Adjusted PR (95% CI) | p value |
| Survey year |  | 1.01 (1.01-1.02) | <0.001 | 1 (0.99-1) | 0.166 | 1.07 (1.06-1.08) | <0.001 | 1.06 (1.05-1.07) | <0.001 | 0.99 (0.99-1.00) | 0.107 | 0.99 (0.99-1) | 0.026 |
| Age (years) |  | 1.01 (1.01-1.01) | <0.001 | † |  | 1.05 (1.05-1.05) | <0.001 | † |  | 1.01 (1.01-1.01) | <0.001 | † |  |
| Sex |  |  |  |  |  |  |  |  |  |  |  |  |  |
|  | Men | Reference |  | Reference |  | Reference |  | Reference |  | Reference |  | Reference |  |
|  | Women | 1.19 (1.17-1.21) | <0.001 | 1.15 (1.13-1.18) | <0.001 | 1.22 (1.14-1.29) | <0.001 | 1.12 (1.05-1.19) | <0.001 | 1.02 (0.98-1.07) | 0.273 | * |  |
| Educational level |  |  |  |  |  |  |  |  |  |  |  |  |  |
|  | Primary | Reference |  | Reference |  | Reference |  | Reference |  | Reference |  | Reference |  |
|  | Secondary | 1.15 (1.12-1.19) | <0.001 | 1.06 (1.03-1.09) | <0.001 | 0.75 (0.70-0.80) | <0.001 | 0.79 (0.74-0.86) | <0.001 | 0.95 (0.90-1.00) | 0.036 | 0.95 (0.9-1) | 0.035 |
|  | Higher | 1.83 (1.78-1.88) | <0.001 | 1.35 (1.31-1.39) | <0.001 | 0.76 (0.71-0.82) | <0.001 | 0.65 (0.6-0.71) | <0.001 | 0.96 (0.92-1.01) | 0.158 | 0.94 (0.89-0.99) | 0.033 |
|  | NK/NR | 0.70 (0.61-0.82) | <0.001 | 0.78 (0.67-0.89) | <0.001 | 0.65 (0.46-0.91) | 0.012 | 1.05 (0.76-1.45) | 0.758 | 0.79 (0.58-1.07) | 0.132 | 0.77 (0.56-1.06) | 0.115 |
| Marital status |  |  |  |  |  |  |  |  |  |  |  |  |  |
|  | Single | Reference |  | Reference |  | Reference |  | Reference |  | Reference |  | Reference |  |
|  | Married | 1.51 (1.46-1.66) | <0.001 | 1.38 (1.34-1.43) | <0.001 | 3.69 (3.25-4.18) | <0.001 | 2.73 (2.4-3.1) | <0.001 | 1.3 (1.17-1.46) | <0.001 | 1.26 (1.13-1.4) | <0.001 |
|  | Cohabiting | 1.00 (0.97-1.04) | 0.838 | 1.12 (1.09-1.16) | <0.001 | 1.6 (1.41-1.82) | <0.001 | 1.6 (1.41-1.82) | <0.001 | 1.09 (0.97-1.22) | 0.151 | 1.09 (0.97-1.22) | 0.136 |
|  | Widowed | 1.53 (1.46-1.60) | <0.001 | 1.53 (1.46-1.6) | <0.001 | 6.49 (5.63-7.47) | <0.001 | 3.3 (2.83-3.85) | <0.001 | 1.38 (1.23-1.55) | <0.001 | 1.3 (1.16-1.46) | <0.001 |
|  | Divorced/separated | 1.32 (1.27-1.37) | <0.001 | 1.27 (1.22-1.32) | <0.001 | 3.7 (3.22-4.25) | <0.001 | 2.67 (2.32-3.08) | <0.001 | 1.21 (1.07-1.36) | 0.002 | 1.18 (1.05-1.32) | 0.006 |
| Wealth index |  |  |  |  |  |  |  |  |  |  |  |  |  |
|  | Poorest | Reference |  | Reference |  | Reference |  | Reference |  | Reference |  | Reference |  |
|  | Poor | 1.6 (1.55-1.66) | <0.001 | 1.48 (1.42-1.54) | <0.001 | 1.91 (1.74-2.10) | <0.001 | 1.75 (1.56-1.96) | <0.001 | 1.21 (1.12-1.30) | <0.001 | 1.14 (1.04-1.24) | 0.003 |
|  | Middle | 2.1 (2.03-2.17) | <0.001 | 1.82 (1.74-1.9) | <0.001 | 2.61 (2.38-2.86) | <0.001 | 2.24 (1.97-2.55) | <0.001 | 1.19 (1.10-1.28) | <0.001 | 1.08 (0.99-1.19) | 0.099 |
|  | Rich | 2.65 (2.57-2.74) | <0.001 | 2.15 (2.05-2.25) | <0.001 | 3.24 (2.95-3.56) | <0.001 | 2.67 (2.33-3.06) | <0.001 | 1.21 (1.12-1.30) | <0.001 | 1.09 (0.99-1.2) | 0.085 |
|  | Richest | 3.51 (3.4-3.62) | <0.001 | 2.51 (2.39-2.64) | <0.001 | 4.05 (3.68-4.44) | <0.001 | 3.14 (2.72-3.61) | <0.001 | 1.34 (1.25-1.44) | <0.001 | 1.19 (1.07-1.31) | 0.001 |
| Health insurance |  |  |  |  |  |  |  |  |  |  |  |  |  |
|  | Uninsured | Reference |  | Reference |  | Reference |  | Reference |  | Reference |  | Reference |  |
|  | Insured | 1.75 (1.7-1.8) | <0.001 | 1.69 (1.64-1.74) | <0.001 | 1.66 (1.53-1.81) | <0.001 | 1.39 (1.27-1.51) | <0.001 | 1.17 (1.09-1.25) | <0.001 | 1.14 (1.06-1.21) | <0.001 |
| Area of residence |  |  |  |  |  |  |  |  |  |  |  |  |  |
|  | Urban | Reference |  | Reference |  | Reference |  | Reference |  | Reference |  | Reference |  |
|  | Rural | 0.47 (0.46-0.48) | <0.001 | 0.92 (0.89-0.96) | <0.001 | 0.37 (0.35-0.40) | <0.001 | 0.76 (0.69-0.84) | <0.001 | 0.83 (0.79-0.88) | <0.001 | 0.93 (0.87-1) | 0.059 |
| Natural region |  |  |  |  |  |  |  |  |  |  |  |  |  |
|  | Coast | Reference |  | Reference |  | Reference |  | Reference |  | Reference |  | Reference |  |
|  | Highlands | 0.6 (0.59-0.62) | <0.001 | 0.86 (0.84-0.88) | <0.001 | 0.46 (0.43-0.49) | <0.001 | 0.74 (0.69-0.8) | <0.001 | 0.79 (0.74-0.83) | <0.001 | 0.82 (0.77-0.87) | <0.001 |
|  | Rainforest | 0.64 (0.62-0.65) | <0.001 | 0.98 (0.95-1) | 0.074 | 0.67 (0.63-0.72) | <0.001 | 1.15 (1.07-1.23) | <0.001 | 0.94 (0.90-0.99) | 0.01 | 0.99 (0.94-1.04) | 0.693 |
| Hypertension |  |  |  |  |  |  |  |  |  |  |  |  |  |
|  | No hypertension | Reference |  | Reference |  | Reference |  | Reference |  | Reference |  | Reference |  |
|  | Hypertension | 1.49 (1.45-1.52) | <0.001 | 1.37 (1.34-1.4) | <0.001 | 4.34 (4.09-4.61) | <0.001 | 3.19 (2.99-3.4) | <0.001 | 1.18 (1.13-1.23) | <0.001 | 1.12 (1.08-1.17) | <0.001 |
| Daily smoker |  |  |  |  |  |  |  |  |  |  |  |  |  |
|  | No | Reference |  | Reference |  | Reference |  | Reference |  | Reference |  | Reference |  |
|  | Yes | 0.93 (0.85-1.02) | 0.128 | 0.88 (0.81-0.95) | 0.002 | 1.30 (1.06-1.59) | 0.011 | 1.21 (0.98-1.47) | 0.07 | 0.84 (0.70-1.01) | 0.063 | 0.86 (0.72-1.02) | 0.09 |
| BMI (kg/m2) |  | 1.03 (1.02-1.03) | <0.001 | † |  | 1.04 (1.03-1.04) | <0.001 | † |  | 1.00 (1.00-1.00) | 0.896 | * |  |

\*: Not included in the adjusted model because it had a p-value >0.20 in the crude model.
†: Not included in the adjusted model due to a VIF >10.
PR: prevalence ratio, NK/NR: Not known/no response, BMI: body mass index

Regarding PDD, in the crude analysis each additional year of age increased prevalence by 5% (p<0.05). PDD was also higher among women and lower among individuals with secondary or higher education compared to those with primary education or less. Being married, widowed, or divorced was associated with higher PDD, with the strongest association among widowed individuals. Higher wealth index and health insurance coverage were associated with higher PDD (p<0.05), while living in rural areas (vs. urban) and in the Highlands (vs. Coast) was associated with lower PDD (p<0.05). PDD was 3.2 times higher among individuals with hypertension (p<0.05). Age and BMI were not included in the adjusted model due to multicollinearity (Table 3).

Among individuals with diabetes, in the crude model PDT increased by 1% per year of age. In the adjusted model, PDT was lower among individuals with secondary or higher education; and higher among married, widowed, or divorced individuals; those from the highest wealth quintile; and those with any type of health insurance. Living in rural areas was associated with lower PDT in the crude model (p<0.05), but became non-significant in the adjusted model (p=0.059). Living in the Highlands was associated with lower PDT (p<0.05), while hypertension with a 12% higher PDT (p<0.05). Age was not included in the adjusted model due to multicollinearity (Table 3).

### Trend Analysis

Temporal trends in screening, diagnosis, and treatment are presented in aggregate in Table 4 and Figure 2 and stratified by population characteristics in Supplementary Figures S1–S10.

**Figure 2.**
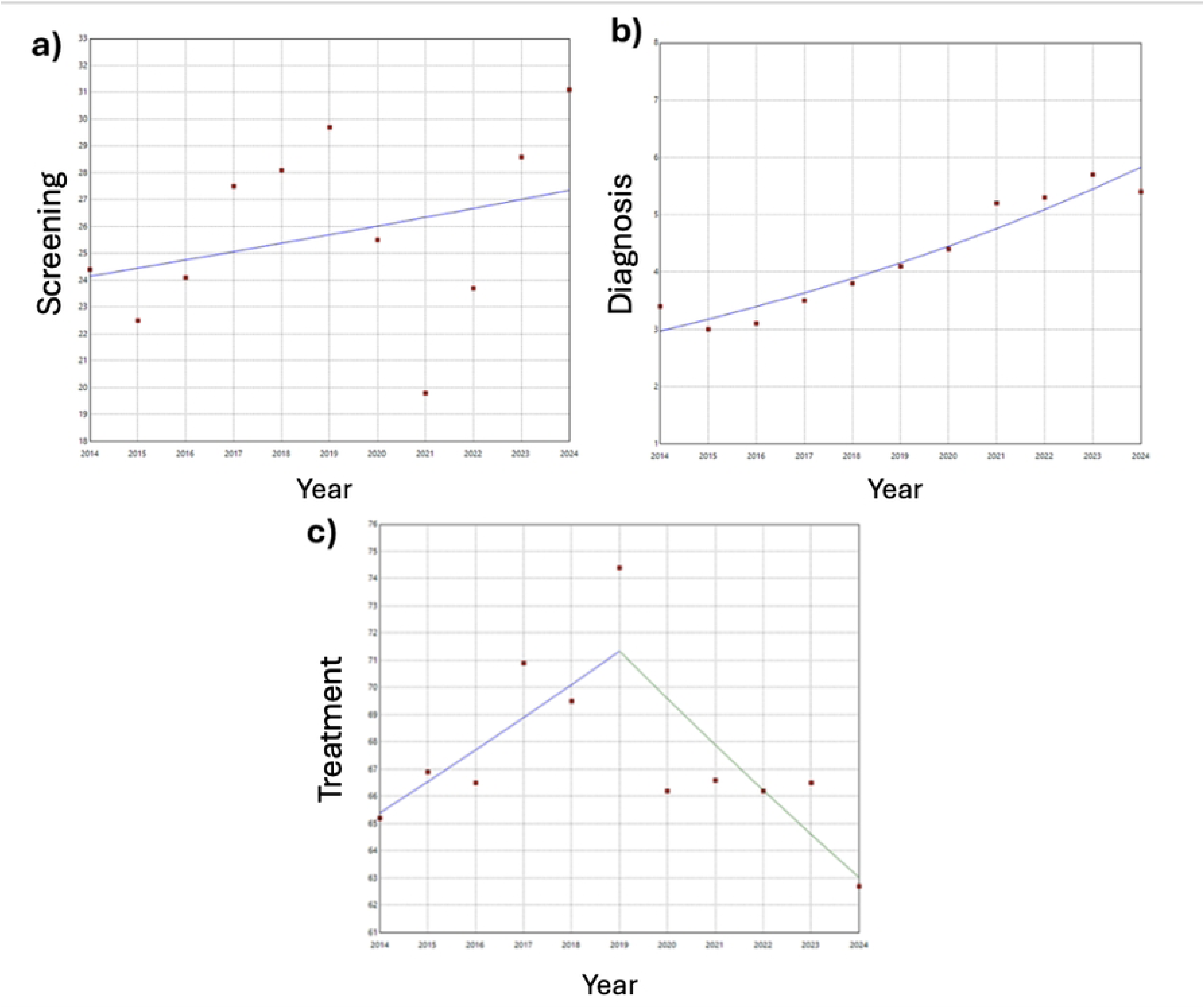
National time trends in a) screening [APC 2014-2024: 1.3 (95%CI: -2,4; 5.0)], b) diagnosis [APC 2014-2024: 7.0 (95%CI: 4.5; 9.5)] and c) treatment [APC 2014-2019: 1.8 (95%CI: 0.3; 6.3) and APC 2019-2024: -2.4 (95%CI: -6.3; -1)], 2014-2024.

**Table 4:**
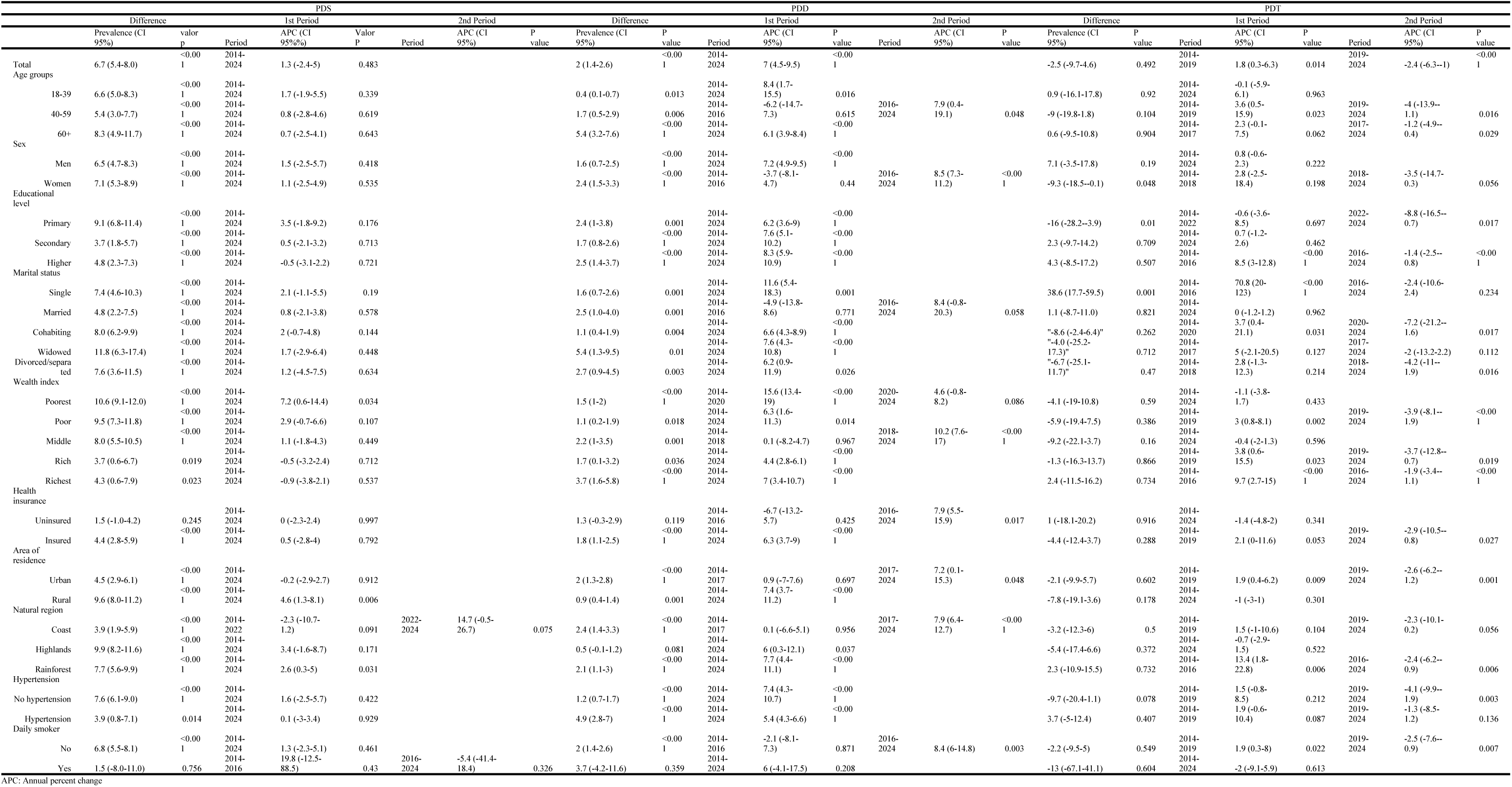
Time trends in PDS, PDD, PDT for diabetes according to associated factors in 2014-2024.

Between 2014 and 2024, the PDS increased by 6.7% (95% CI: 5.4–8.0), although the APC was not statistically significant (1.3%) (Table 4). When stratified by age group, sex, educational level, marital status, insurance status, daily smoking, and hypertension, PDS showed a general increasing pattern—except among individuals with higher education, the uninsured, and smokers (after 2016)—yet APC estimates were not significant.

Two distinct patterns emerged by wealth index. While the poorest quintile showed a sustained annual increase (APC: 7.2%), the richest quintile exhibited a non-significant decrease (APC: –0.9%). By area of residence, PDS significantly increased only in rural areas (APC: 4.6%), with no significant changes observed in urban areas (APC: –0.2%). By natural region, the Highlands (APC: 3.4%) and Rainforest (APC: 2.6%) displayed a single increasing pattern, whereas the Coast presented two segments: a non-significant decrease (APC 2014–2022: –2.3%) followed by a significant increase (APC 2022–2024: 14.7%) (Table 4).

The PDD increased by 2.0 percentage points (95% CI: 1.4–2.6%), with a significant average annual increase of 7.0% over 2014–2024; this increase was observed across most subgroups. Among men, the upward trend was sustained (APC: 7.2%), while among women the trend became significant beginning in 2016 (APC: 8.5%). When stratified by age group, educational level, marital status, daily smoking, and hypertension, a general increasing trend was observed, except among those aged 40–59 years, married individuals, and non-smokers, who showed an initial non-significant decrease followed by a increase (significant for those aged 40–59 years and non-smokers) (Table 4).

All wealth quintiles exhibited a single increasing pattern in PDD except for the poorest and richest quintiles, which showed two increasing segments. Unlike insured individuals—who experienced a single significant increase (APC: 6.3%) in PDD from 2014 to 2024—the uninsured displayed an initial non-significant decrease (APC 2014–2016: –6.7%) followed by a significant increase (APC 2016–2024: 7.9%). The upward trend in PDD from 2014 to 2024 was also observed when stratifying by area of residence and natural region, though trends split into two segments in urban areas and in the Coast (Table 4).

Between 2014 and 2024, PDT decreased by 2.5%, with an initial significant increase (APC 2014–2019: 1.8%) followed by a significant decrease (APC 2019–2024: –2.4%). When stratified by age group, an initial increase followed by a decline was observed among those aged 40–59 years (APC 2014–2019: 3.6%; APC 2019–2024: –4.0%) and ≥60 years (APC 2014–2017: 2.3%; APC 2017–2024: –1.2%). This pattern also appeared among women (APC 2014–2018: 2.8%; APC 2018–2024: –3.5%) and among those with higher education (APC 2014–2016: 8.5%; APC 2016–2024: –1.4%), whereas individuals with primary education or less showed two distinct periods of decline (APC 2014–2022: –0.6%; APC 2022–2024: –8.8%) (Table 4).

By marital status, with the exception of married individuals—who showed no significant changes—all categories displayed an initial increase, strongest among single individuals (APC 2014–2016: 70.8%), followed by a subsequent decrease. While the poorest quintile experienced a single decreasing pattern (APC: –1.1%), the richest quintile displayed an initial significant increase (APC 2014–2016: 9.7%) followed by a significant decrease (APC 2016–2024: –1.9%). Similarly, an initial increase followed by a decrease was observed among insured individuals (APC 2014–2019: 2.1%; APC 2019–2024: –2.9%), those living in urban areas (APC 2014–2019: 1.9%; APC 2019–2024: –2.6%), residents of the Coast (APC 2014–2019: 1.5%; APC 2019–2024: –2.3%), the Rainforest (APC 2014–2016: 13.4%; APC 2016–2024: –2.4%), and among non-smokers (APC 2014–2019: 1.9%; APC 2019–2024: –2.5%) (Table 4).

## DISCUSSION

### Main findings

In this study, we used nationally representative data to assess trends and factors associated with diabetes-related healthcare utilization in Peru over the past 11 years.

The PDS increased from 24% to 31% between 2014 and 2024, and was higher among adults ≥60 years, women, individuals with higher education, those who were married or widowed, those in higher wealth quintiles, retirees, insured individuals, residents of urban areas and the Coast, as well as those with hypertension and higher BMI. Factors associated with higher PDS included being female, having higher education, not being single, higher wealth, being insured, and having hypertension. Conversely, living in rural areas and in the Highlands was associated with lower PDS. In the trend analysis, despite the decline observed in 2020–2021, we found a sustained overall increase, reaching levels in 2024 that exceeded those observed before the pandemic.

Between 2014 and 2024, the PDD increased from 3% to 5%, with higher values observed among adults ≥60 years, women, individuals with higher education, widowed individuals, retirees, those in higher wealth quintiles, the insured, residents of urban areas and the Coast, individuals with hypertension, and those with higher BMI. Factors associated with higher diagnosis included being female, not being single, higher wealth, being insured, living in the Rainforest, and having hypertension. In contrast, secondary or higher education, as well as living in rural areas and in the Highlands, were associated with lower diagnosis. The trend analysis showed a sustained annual increase in PDD throughout the study period.

The PDT decreased from 65% to 63% between 2014 and 2024, with higher values observed among adults ≥60 years, men from 2020 onward, retirees, married or widowed individuals, the insured, residents of urban areas and the Coast, and those with hypertension. Factors associated with higher PDT included not being single, belonging to the richest wealth quintile, being insured, and having hypertension. In contrast, higher education and living in the Highlands were associated with lower treatment. Regarding trends, PDT showed a sustained increase until 2019, followed by a consistent decline during the latter half of the period, reaching its lowest level in 2024.

### Results in context

There is heterogeneity in the literature regarding PDS. The 31% value found for 2024 in our study is slightly higher than the rate reported in Mexico in 2023 [30], but lower than the 42% identified between 2005–2021 in the United States, according to National Health and Nutrition Examination Survey (NHANES) data [31]. These differences may be attributed to the broader healthcare coverage in the United States and to the fact that NHANES assesses diabetes screening within the past three years—a longer interval than the one used in ENDES [31].

The 2024 PDD of 5% in our study was lower than the 12% reported in adults ≥20 years in Mexico in 2023 and the 8% among adults aged 20–79 years in the United States in 2012 [30,32]. These differences may be due to a lower actual prevalence of diabetes (i.e., diagnosed plus undiagnosed) in the Peruvian population [33,34] as well as lower access to screening services [31]. On the other hand, our 2023 results (6%) are similar to preliminary findings by Fiestas-Saldarriaga et al., who reported a 6% PDD in Peruvians ≥15 years using ENDES 2023 [35].

The national PDT was 63% in 2024. This value is lower than the 78% observed among people with diabetes in Mexico [30]. In a study evaluating 28 countries using national survey data from LMICs, our result was lower than the 74% reported in Costa Rica in 2010, comparable to the 65% in Guyana in 2016, and higher than the 48% in Chile in 2009–2010 [8]. These differences may be attributed to inequities in health system access and medication availability, as well as methodological differences across surveys [8].

Consistent with other studies [12,20,31], we found that older age is associated with higher PDS in the previous 12 months; this can be explained by increased contact with health services for preventive examinations as people age, as well as the higher risk of diabetes that may prompt clinicians to request screening [15,20].

We observed that women had higher PDS, which aligns with studies conducted in the United States between 1976–1989 and 2016–2019 [15,16]. This may be attributed to the fact that women use preventive services more frequently than men [36]. Higher educational attainment, greater wealth, and health insurance were associated with higher PDS, consistent with previous evidence [12,16,31]. These associations may be explained by greater health literacy among more educated individuals and increased ability to access and pay for screening tests among those with insurance or higher wealth [12].

In our study, living in urban areas or on the Coast was associated with higher PDS, likely due to better access to and use of outpatient care services [37]. Hypertension was also associated with higher screening, consistent with various studies, and can be attributed to guideline recommendations to screen individuals with hypertension due to their higher diabetes risk [15,16,31,38].

Older age was associated with higher PDD, consistent with the findings of Malta et al. in Brazil [17]. This can be explained by the greater diabetes risk associated with aging [39] as well as more frequent screening. Being a woman, belonging to higher wealth quintiles, and having health insurance were also associated with higher PDD, likely reflecting higher screening rates in these groups. We found an inverse association between educational attainment and PDD, consistent with previous studies [17,40,41]. This may be explained by low health literacy among individuals with lower education, which may lead to behaviours that increase diabetes risk [41,42].

Being single was associated with lower PDD, potentially due to differences in the diabetes risk profile (e.g., younger age). Similarly, living in rural areas and in the Highlands was associated with lower PDD, which correlates with lower screening frequency in these areas. As in the 2019 study of the Brazilian population [17], having hypertension was associated with higher PDD, likely due to an increased diabetes risk in this group [38] as well as more frequent screening.

We found that higher educational attainment was associated with lower PDT, which contrasts with previous studies [8,43]. This finding may reflect differences in self-reporting of treatment or that individuals with higher education achieve better glycemic control and require less medication. We also found that being single was associated with lower PDT compared to other marital statuses, consistent with the study by Vieira and Santos [44], and may be explained by the role of family support as a facilitator of disease management [45].

On the other hand, belonging to the richest quintile compared to the poorest was associated with higher treatment. This aligns with findings from a systematic review [22] (39), which identified medication cost as a barrier to treatment, especially among lower-income groups [21,22]. Compared with the Coast, living in the Highlands was associated with lower treatment, independent of wealth and education. This may be due to poorer access to health services in this region, limiting attendance to follow-up visits and medication collection. Similarly, having health insurance was associated with higher PDT, consistent with findings from the 2014–2019 Peruvian population study by Calderon-Ramirez et al. [24].

In this study, we observed a 27% increase in PDS between 2014 and 2024 (11 years), with an overall upward trend during the study period. This finding is comparable to results reported among adults ≥20 years in Ontario, Canada between 1995–2005, where fasting plasma glucose PDS increased by 28% [46]. These increases may be explained by rising metabolic risk factors in the Peruvian population (e.g., obesity) [47], expansion of health service access driven by universal health coverage strategies [48], and the dissemination and gradual adoption of national clinical guidelines recommending diabetes screening [49].

We observed a sustained annual increase in PDD between 2014 and 2024. This pattern is similar to that reported by Geiss et al., who observed a 5% APC in adults aged 20–79 years in the United States during 1990–2008 using National Health Interview Survey data [32]. This increase could be attributed to both a real rise in diabetes prevalence driven by greater exposure to metabolic risk factors [47] and population aging [50], as well as greater case detection resulting from increased PDS.

Despite an initial sustained increase in PDT, we observed a decline over the past five years after 2020. The initial increase is consistent with global and Latin American patterns reported by the NCD Risk Factor Collaboration, where most countries have shown increasing treatment coverage from 1990 to 2022 [4]. This improvement may be explained by greater access to health services and medications [48], increased case detection, and guideline implementation that emphasizes pharmacological management of diabetes [49].

Conversely, the decline in treatment access during 2020–2021 may be explained by the impact of the COVID-19 pandemic, which reduced access to outpatient consultations and hindered medication collection, in addition to causing unemployment and financial difficulties that affected the ability to afford medications [51].

### Strengths and limitations

As we used a nationally representative survey, our findings can be generalized to the adult population in Peru. Furthermore, because the ENDES has been conducted annually since 2014, we were able to examine 11-year trends in three key indicators of diabetes-related health services: screening, diagnosis and treatment. Therefore, our study is well positioned to inform clinicians, policymakers and other relevant stakeholders seeking to improve diabetes care in Peru.

Nonetheless, this study presents limitations that should be considered when interpreting the results. First, by using secondary data, the analysis was limited to the variables available and it was not possible to obtain additional information, as the records were anonymized.

Diabetes variables were based on self-report and did not include biomarkers such as fasting plasma glucose or glycated hemoglobin. Although these indicators would provide additional epidemiological information, their absence does not affect the estimation of access to health services—the primary objective of our study.

The use of self-report in ENDES for measuring diabetes variables may introduce potential information bias due to recall bias or social desirability. This limitation is inherent to the study design and may have led to slight under- or over-estimations of each diabetes indicator. However, INEI employs trained and experienced interviewers to administer the questionnaires [14].

Finally, because this was a cross-sectional analysis, it is not possible to establish causal relationships between the factors studied and diabetes screening, diagnosis, or treatment. Rather, we explored broad patterns to provide an overview of potential factors related to access to diabetes screening, diagnosis, and treatment in the Peruvian context.

### Conclusions

Over the past 11 years, PDS and PDD showed an overall increasing trend, whereas PDT demonstrated a decline in the last five years. Additionally, variations in access to these three services were observed according to sociodemographic and clinical characteristics. These findings highlight the need to consider these differences in the planning and organization of diabetes-related health services in Peru.

## Contributions

WCG-V: conceptualization, methodology, software, analysis, investigation, data curation, writing – original draft, visualization. MC-R: conceptualization, writing – review and editing, project administration. GS-S: conceptualization, methodology, software, analysis, investigation, writing – original draft, writing – review and editing, supervision, project administration. RT-R: conceptualization, writing – review and editing, project administration.

## Data availability

We used nationally-representative survey data that are in the public domain (https://proyectos.inei.gob.pe/microdatos/), which were accessed on September 19^th^, 2025. We provide the code of data preparation and data analysis as supplementary materials.

